# Cardiac involvement in Fabry disease: Implications of retinal vessel analysis in detecting early microvascular alterations despite ERT

**DOI:** 10.1101/2025.01.14.25320513

**Authors:** Timon Kuchler, Claudia Regenbogen, Roman Günthner, Andrea Reiberio, Javier Carbajo, Nora Hannane, Michael Wunderle, Abdalrahman Assaf, Maciej Lech, Henner Hannsen, Lukas Streese, Derralynn Hughes, Bernhard Haller, Konstantin Kotliar, Uwe Heemann, Christoph Schmaderer

## Abstract

**Background:** Cardiac complications driven by microvascular changes are one of the primary reasons for mortality in patients with Fabry disease (FD). While enzyme replacement therapy (ERT) can effectively clear globotriaosylceramide (Gb3) deposits in the endothelium, it remains controversial whether ERT fully resolves microvascular dysfunction, particularly in advanced cases.

**Methods:** We conducted a cross-sectional observational study of 63 FD patients, of whom 60 had quality retinal vessel analysis (RVA) data and were age- and gender-matched to a healthy control group (HC, n=60, matched out of 204). Associations with disease severity, echocardiographic and laboratory parameters, and ERT were explored.

**Results:** FD patients exhibited significantly reduced venular flicker-induced dilation (vFID, 3.5% ± 1.6% vs. 4.6% ± 2.4%; p = 0.006), narrower central retinal arteriolar equivalents (CRAE, 165.2 µm [153.5 - 183.2] vs. 183.2 µm [174.7 - 191.4], p < 0.001), and lower arteriolar-venular ratio (AVR, 0.82 [0.74 - 0.87] vs. 0.86 [0.82 - 0.91]; p < 0.001) compared to HC, independent of cardiovascular risk factors. Despite ERT, retinal microvascular health remains incompletely recovered in severely affected FD patients. Narrower retinal arterioles are closely associated with echocardiographic parameters and laboratory markers of cardiac involvement. Additionally, CRAE demonstrated accuracy comparable to LysoGb3 levels in identifying GLA variants with a potential cardiac phenotype. FD patients exhibited elevated chronic inflammation and endothelial dysfunction markers, including Rantes, MCP1, CXCL10, ICAM-1, VCAM-1, and VEGF. In FD patients with cardiac variants, higher levels of inflammatory parameters were associated with impaired retinal microcirculation.

**Conclusion:** Our findings demonstrate that RVA can detect microvascular alterations in FD, offering a potential non-invasive approach for assessing disease severity and monitoring cardiovascular health. The persistence of retinal microvascular changes despite ERT may reflect the need for earlier interventions. Longitudinal studies are warranted to further explore the predictive value of RVA for Fabry-related cardiovascular outcomes.

**Registration:** https://clinicaltrials.gov/study/NCT06758648; Unique identifier: NCT06758648.

**Novelty and Significance:** What Is Known?

- Fabry disease (FD) is an X-linked lysosomal storage disorder characterized by the accumulation of globotriaosylceramide (Gb3), leading to microvascular dysfunction and cardiac complications.
- Enzyme replacement therapy (ERT) and pharmacological chaperone therapy (PCT) are available treatments, but their ability to fully reverse microvascular changes remains uncertain.
- Retinal vessel analysis (RVA) is a non-invasive tool previously used to monitor systemic microvascular health in cardiovascular diseases.

What New Information Does This Article Contribute?

- Retinal microvascular abnormalities, including arteriolar narrowing and reduced venular dilation, are detectable in FD, even in patients receiving ERT.
- Retinal vessel parameters are strongly associated with systemic markers of cardiac involvement.
- RVA offers a non-invasive, cost-effective method to assess microvascular health and could serve as a valuable tool for disease monitoring and stratification in FD.

**Summary:** FD is a rare genetic disorder associated with significant microvascular and cardiac complications due to the progressive accumulation of Gb3. Although ERT is a cornerstone of FD management, we demonstrate that retinal microvascular abnormalities persist despite treatment, particularly in patients with advanced disease. By employing RVA, the study highlights the close association between retinal arteriolar narrowing, systemic markers of cardiac involvement and disease severity. These findings underscore the potential of RVA as a non-invasive, cost-effective method for assessing endothelial health, monitoring disease severity and cardiac involvment in FD. Additionally, the results suggest that earlier interventions and adjunctive therapies targeting endothelial dysfunction and inflammation may be necessary to improve outcomes in advanced cases. RVA could play a pivotal role in routine care by providing insights into systemic vascular health and optimizing Fabry-specific therapies.

## Introduction

Fabry disease (FD, OMIM-301500) is a rare, X-linked lysosomal storage disorder with a reduced or completely absent activity of the enzyme α-galactosidase A encoded by the GLA-gene. This malfunction leads to a progressive accumulation of glycosphingolipids, such as globotriaosylceramide (Gb3), in the endothelium of different organs. Primary cellular dysfunction and secondary inflammation result in reduced organ perfusion and fibrosis, which causes progressive, multisystemic organ damage [1–3]. In adult FD patients, cardiovascular (CV) manifestations, including left ventricular hypertrophy (LVH), and cerebrovascular complications, including stroke, contribute to a significantly lower life expectancy [4–6]. Heterozygous females with α-galactosidase A deficiency may also develop Fabry-associated complications, although usually with a slower progression [7]. Diagnostic delay of FD can reduce the efficacy of the treatment, and cardiovascular complications may significantly benefit from early treatment [5, 8, 9].

Although the exact pathophysiological mechanisms underlying endothelial dysfunction (ED) in FD remain unclear, Gb3 has been shown to induce the degradation of endothelial K_Ca_ 3.1 channels, which play a crucial role in endothelium-dependent hyperpolarization. Additionally, secondary chronic inflammation may contribute to the degradation of the endothelial glycocalyx, further exacerbating ED in FD patients [10, 11]. Consequently, various markers of endothelial activation and chronic inflammation, including vascular endothelial growth factor (VEGF), intercellular adhesion molecule-1 (ICAM-1), and monocyte chemoattractant protein-1 (MCP1), are elevated in patients with Fabry disease [12–15].

After the administration of enzyme replacement therapy (ERT), complete microvascular clearance of Gb3 in the endothelium and podocytes has been observed. However, studies indicate that Gb3 clearance does not prevent altered molecular signaling, further underscoring the importance of early disease detection [16, 17].

Quantifying ED by non-invasive imaging methods has been proposed by the European Society of Cardiology (ESC) [18], and studies have shown impaired endothelial function in medium-to large-sized vessels via pulse-wave velocity (PWV) and flow-mediated dilation (FMD) [19, 20]. However, cardiovascular complications in FD are mainly a result of coronary microvascular dysfunction, and monitoring the microcirculation with non-invasive tools could potentially provide a better tool for early diagnosis and disease monitoring [16, 21, 22]. To our knowledge, data on small vessel health in FD patients remains limited.

Dynamic retinal vessel analysis (DVA) and static retinal vessel analysis (SVA) are two quantitative and non-invasive technologies that follow up on changes in retinal microcirculation as a predictive tool for the progression of systemic cardiovascular disease (CVD). DVA quantifies the reaction to flickering light over time, mainly mediated by neuro-vascular coupling followed by shear-stress-induced nitric oxide (NO) release [23]. This allows direct assessment of the cerebrovascular function, and especially in CVD, DVA and SVA have shown in multiple studies their high potential to monitor endothelial health non-invasively [24, 25]. The Atherosclerosis Risk in Communities (ARIC) study proved the excellent predictive value of retinal arteriolar narrowing and venular widening as a long-term predictor of CV events [26]. In a large group of dialysis patients, we were able to show that impaired retinal venular dilation (vFID) is an independent predictor of all-cause mortality [27].

Early detection of organ damage in FD is crucial, however diagnosis in early stages still presents a considerable challenge. Therefore, the study’s primary aim was to investigate whether the retinal structure and function, measured by DVA and SVA, are altered in FD. The secondary aim was to evaluate how parameters of the retinal microcirculation are associated with clinical disease severity, enzyme replacement therapy (ERT), and the cardiac phenotype.

## Methods

### Study design and patients

This study is part of an observational longitudinal investigation examining the retinal microvasculature of 63 FD patients and providing an in-depth clinical characterization of FD patients. All patients were recruited in our outpatient clinic from the department of nephrology (Klinikum Rechts Der Isar, Technical University of Munich, School of Medicine), which all underwent clinical examination, RVA analysis, and blood collection. The local ethics committee approved the study protocol (Ethics Committee of the Klinikum Rechts Der Isar, Technical University of Munich, School of Medicine, No. 432/19S). The study “VASCinFABRY” was registered previously at ClinicalTrials.gov (NCT06758648). All participants of this study gave written informed consent. The study conforms to the principles of the Declaration of Helsinki and was designed following the Strengthening the Reporting of Observational Studies in Epidemiology (STROBE) guidelines. The sample size was defined previously in the study protocol and was powered for the primary aim of comparing retinal vessel parameters in Fabry patients with a healthy cohort (HC). Assumptions regarding expected differences and variability were informed by comparable patient cohorts, and the calculation was conducted with guidance from a statistical expert. The HC was recruited as described previously [28].

Inclusion criteria were age >18, diagnosis of FD by genetic testing of the galactosidase alpha (GLA) gene, α-gal A activity in leukocytes, or elevated LysoGb3 levels. Exclusion criteria were active infection and or malignant disease, operation two weeks before baseline examination, known glaucoma or epilepsy, and lack of written consent. Genetic analysis was done in 62 patients (Genetic analysis, Molecular Genetics and Metabolism Laboratory, Centogene AG, Rostock, Germany).

### Retinal Vessel Analysis

DVA and SVA were performed by experienced and trained examiners using the Dynamic Retinal Vessel Analyzer (DVAlight; IMEDOS Systems, Jena, Germany) and the Static Vessel Analyzer (IMEDOS Systems GmbH, Jena, Germany). The technique has been extensively described in our studies [27, 29]. SVA was performed before DVA. Before the examination, pupils were dilated using topical tropicamide (0.5% Mydriaticum Stulln; Pharma Stulln, Germany), and patients were seated in a quiet, dark room for a ten-minute rest period.

In the case of DVA, patients were asked to focus on a needle attached inside the camera, and one arteriole and venule diameter were automatically and continuously recorded for 350 seconds. Arteriole and venule segments between 0.5 to 1 mm were analyzed approximately one disc diameter away from the optic nerve in the upper-temporal or lower-temporal direction. The baseline recording was 50 seconds, followed by a flickering phase of 20 seconds and then a recovery period of 80 seconds. Three of these cycles were performed, and we calculated the percentage of maximum arteriolar (aFID) and venular dilation (vFID) to baseline as previously described. For data validation, the quality of vessel response curves was compared using a cumulative scoring method (range 0 to 5) [27]. Retinal measurements with a score <2.5 were excluded after re-evaluation by an experienced observer. In five cases (7.9%), we could not measure vFID due to a lack of information in the measured region, and in six cases (9.5%), we could not obtain high-quality measurements of aFID.

SVA pictures were analyzed using Vesselmap 2^®^ (IMEDOS Systems GmbH, Jena, Germany). One eye was examined, and three images were taken with the focus on the optic disc at an angle of 50°. Roughly one disc diameter away from the optic disc, retinal veins and arterioles segments were semi-automatically labeled. The Paar-Hubbard formula averaged the central retinal arteriolar (CRAE) and central venular (CRVE) equivalents [30]. SVA parameters CRAE and CRVE were measured in measurement units (MU), where 1 MU corresponds to 1 µm in the Gullstrand’s eye model. Inter- and intra-observer inter-class correlation coefficients for CRAE and CRVE range from 0.75 to 0.87 [30, 31]. In three patients (4.8%), we could not obtain high-quality static retinal pictures.

### Analysis of clinical and laboratory characteristics

We studied the medical records to record baseline characteristics, comorbidities, disease onset, gene variants, FD-specific treatment, and medication. Blood examinations and echocardiography took place within the framework of routine tests on the day of baseline recruitment.

Blood samples were collected following previously described protocols [29], and routine parameters were measured in an ISO-certified laboratory. Levels of IL-6, Rantes, VEGF, CXCL10, MCP1, VCAM-1, and ICAM-1 in patient serum were quantified using the Cytometric Bead Array Flex system (BD Biosciences, San Diego, USA), following the manufacturer’s instructions.

The Fabry disease severity scoring system (ADS3) is a validated score that takes four clinical domains (Renal, peripheral nervous system, cardiac, and central nervous system) and one patient-reported outcome (PROM) into account. The average scoring system ranges from “minimal disease severity” (0 p.) to “maximum disease severity” (32 p.) [32]. Interventricular-septum (IVS) thickness was measured and considered as increased for values >13 mm. The presence of chronic kidney disease (CKD) was defined as an eGFR of less than 60ml/min calculated with the CKD-EPI equitation.

In 62 patients, the galactosidase alpha (GLA)-gene sequence variants were analyzed. Variants were classified according to the American College of Medical Genetics and Genomics (ACMG) guidelines, and 33 patients were classified with a pathogenic variant, 7 with a likely pathogenic variant, 8 patients with a variant of uncertain significance (VUS) and 14 with a likely benign variant (**Supplementary Table 1**) [33].

### Statistical Analysis

All statistical analyses were performed using R (4.2.1) and R Studio (Version: 2024.04.2+764). Graphical preparation was done using Affinity Publisher (Version 2.5.5). Normally distributed values are shown as mean, ± standard deviation (SD), and non-normally distributed values as mean and their interquartile range (IQR) if not otherwise stated. To compare patients’ characteristics student’s t-test was used for normally distributed values, the Wilcoxon rank sum test for non-normally, and χ^2^ – test for categorical variables. For comparisons of more than two groups, one-way ANOVA was used for normally distributed data, and Kruskal Wallis was used elsewhere. Normality was assessed with the Shapiro-Wilks test and visually by using histograms. All statistical hypothesis testing was conducted on two-sided 5% significance levels. To match FD patients with the healthy cohort based on age and gender, we used the Matching package; matching success was controlled with the MatchBalance function. Violin- and boxplots were created using the ggplot package. Receiver operating characteristics (ROC) analysis was done with the plotROC and pROC package, and the area under the curve (AUC) was calculated using the trapezoidal rule. The 95% confidence interval (CI) of the AUC was calculated using DeLonǵs method. For the correlation plots and the calculation of Spearmańs (for skewed data) and Pearsońs (for normal data) correlation coefficients, we used the ggscatter function, which is part of the ggpubr package. The correlation heatmap was created using the corplot package. In a multivariable linear model, we adjusted for potential confounders (BMI, arterial hypertension, hypercholesterolemia, and nicotine abuse) of retinal vessel analysis [34–37]. We used the interaction package to create interaction plots. To calculate the p_interaction,_ we fitted a multivariable linear regression model with SVA parameters as the dependent variables, inflammatory and ED parameters, and FD with a potential cardiac variant as two predictor variables (**Supplementary Table 2**). Two independent researchers typed in all clinical data for double-data verification. Efforts were made to minimize missing data throughout the study, and the extent of missingness was kept to a reasonably achievable minimum. Missing data were not imputed. One author had full access to all the data in the study and took responsibility for its integrity and the data analysis.

## Results

### Patients’ characteristics and subgroup analysis

Out of 63 FD patients, 60 (95.2%) had high-quality SVA images (68.3 % female, mean age 48.6 ±16.5 years) and were age- and gender-matched with 60 healthy volunteers (68.3 % female, 49.7 ± 16.2 years) from a previously recruited HC (n = 204) [28]. The occurrence of CV risk factors was comparable between cohorts after matching. There was a tendency towards more nicotine abuse in FD patients; however, not significantly. The most frequent CV risk factors in FD patients were hypercholesterolemia (53.3 %) and obesity (36.7 %). Fabry-related complications included patients with left ventricular hypertrophy (LVH, 48.3 %), patients with central nervous, vascular disease (CNVD, 25.0 %), patients with cardiac arrhythmia (21.7 %), and patients with chronic kidney disease (CKD, 6.7 %). 41 patients (68.3 %) were pre-treated with enzyme replacement therapy (ERT) with a median therapy duration of 1.1 years (0.0 – 3.4). The average Fabry disease severity scoring system (ADS3) was 8.7 (5.2 – 11.8), and median LysoGb3 levels were 4.3 ng/ml (0.9 – 8.3) under current ERT. Laboratory parameters showed significantly higher leukocytes and lower hemoglobin levels in FD patients (**Table 1**).

**Table 1.**
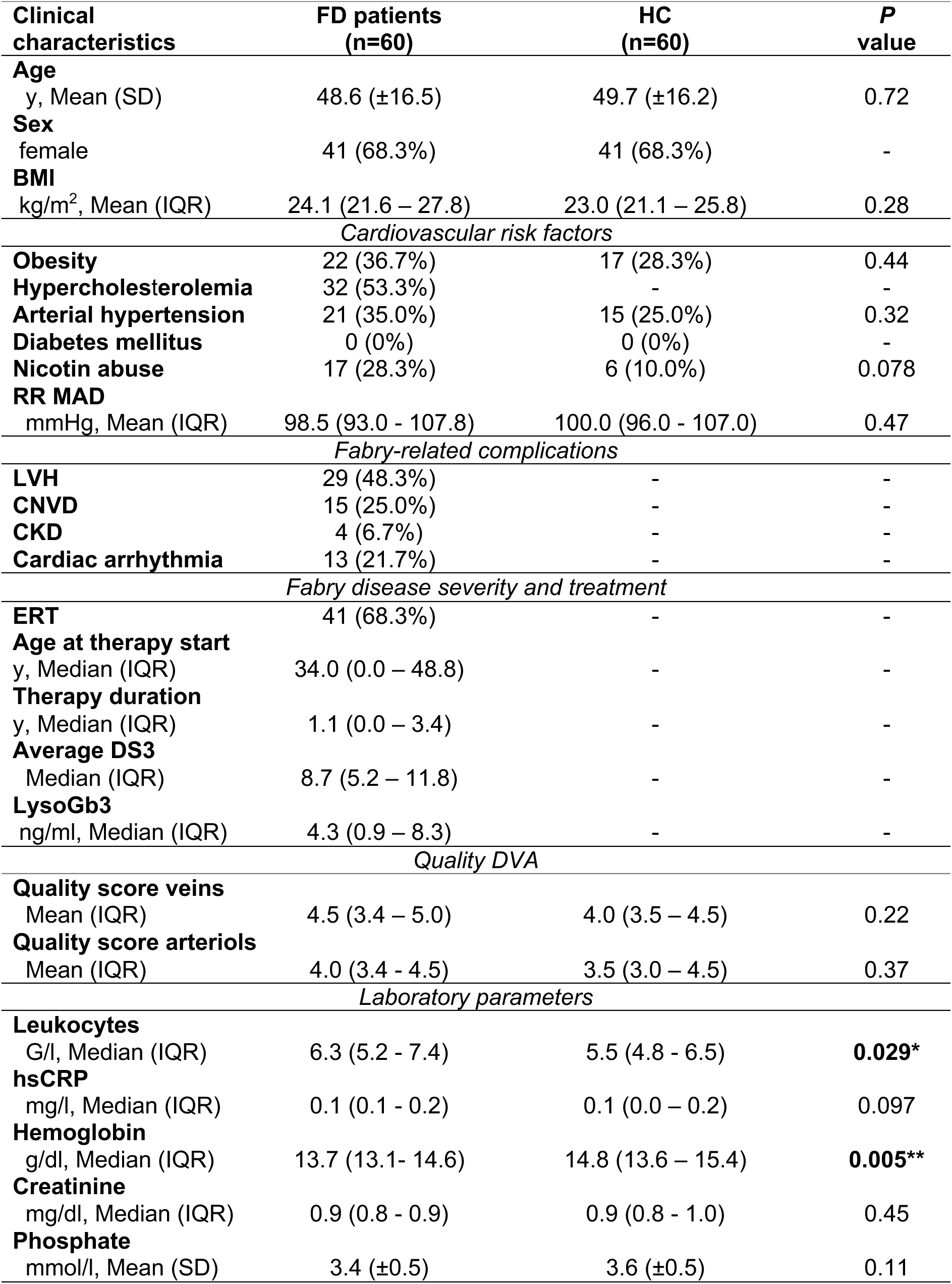
Characteristics of Fabry disease patients compared to age- and gender-matched healthy controls. P-values are shown for statistical tests comparing patients with Fabry disease (FD) (n=60) with an age- and gender matched healthy cohort (HC) (n=60); t-test was used for normally distributed variables, the χ^2^ test for categorical variables, the Wilcoxon rank sum test for variables with a skewed distribution, and Fisher’s exact test for binary variables. BMI, body mass index; RR MAD, the mean arterial pressure; DM, diabetes mellitus I or II; obesity is defined as BMI > 25 kg/m^2^; hypercholesterolemia is defined as cholesterol > 200 mg/dl. LVH, left ventricular hypertrophy is defined as intraventricular thickness > 13 mm; CNVD, central nervous vascular disease is defined as transient ischemic attack (TIA) or Stroke; ERT, Enzyme replacement therapy; DVA, dynamic vessel analysis; Leukocytes (n=100), Hemoglobin (n=99), Creatinine and Phosphate (both n=103) and high-sensitivity C-reactive protein (hs-CRP, n=97) were measured in our cohort.

| **Clinical characteristics** | **FD patients (n=60)** | **HC (n=60)** | **P value** |
| --- | --- | --- | --- |
| **Age** |  |  |  |
| y, Mean (SD) | 48.6 (±16.5) | 49.7 (±16.2) | 0.72 |
| **Sex** |  |  |  |
| female | 41 (68.3%) | 41 (68.3%) | - |
| **BMI** |  |  |  |
| kg/m<sup>2</sup>, Mean (IQR) | 24.1 (21.6 – 27.8) | 23.0 (21.1 – 25.8) | 0.28 |
| *Cardiovascular risk factors* |  |  |  |
| **Obesity** | 22 (36.7%) | 17 (28.3%) | 0.44 |
| **Hypercholesterolemia** | 32 (53.3%) | - | - |
| **Arterial hypertension** | 21 (35.0%) | 15 (25.0%) | 0.32 |
| **Diabetes mellitus** | 0 (0%) | 0 (0%) | - |
| **Nicotin abuse** | 17 (28.3%) | 6 (10.0%) | 0.078 |
| **RR MAD** |  |  |  |
| mmHg, Mean (IQR) | 98.5 (93.0 - 107.8) | 100.0 (96.0 - 107.0) | 0.47 |
| *Fabry-related complications* |  |  |  |
| **LVH** | 29 (48.3%) | - | - |
| **CNVD** | 15 (25.0%) | - | - |
| **CKD** | 4 (6.7%) | - | - |
| **Cardiac arrhythmia** | 13 (21.7%) | - | - |
| *Fabry disease severity and treatment* |  |  |  |
| **ERT** | 41 (68.3%) | - | - |
| **Age at therapy start** |  |  |  |
| y, Median (IQR) | 34.0 (0.0 – 48.8) | - | - |
| **Therapy duration** |  |  |  |
| y, Median (IQR) | 1.1 (0.0 – 3.4) | - | - |
| **Average DS3** |  |  |  |
| Median (IQR) | 8.7 (5.2 – 11.8) | - | - |
| **LysoGb3** |  |  |  |
| ng/ml, Median (IQR) | 4.3 (0.9 – 8.3) | - | - |
| *Quality DVA* |  |  |  |
| **Quality score veins** |  |  |  |
| Mean (IQR) | 4.5 (3.4 – 5.0) | 4.0 (3.5 – 4.5) | 0.22 |
| **Quality score arteriols** |  |  |  |
| Mean (IQR) | 4.0 (3.4 - 4.5) | 3.5 (3.0 – 4.5) | 0.37 |
| *Laboratory parameters* |  |  |  |
| **Leukocytes** |  |  |  |
| G/l, Median (IQR) | 6.3 (5.2 - 7.4) | 5.5 (4.8 - 6.5) | **0.029\*** |
| **hsCRP** |  |  |  |
| mg/l, Median (IQR) | 0.1 (0.1 - 0.2) | 0.1 (0.0 – 0.2) | 0.097 |
| **Hemoglobin** |  |  |  |
| g/dl, Median (IQR) | 13.7 (13.1- 14.6) | 14.8 (13.6 – 15.4) | **0.005\*\*** |
| **Creatinine** |  |  |  |
| mg/dl, Median (IQR) | 0.9 (0.8 - 0.9) | 0.9 (0.8 - 1.0) | 0.45 |
| **Phosphate** |  |  |  |
| mmol/l, Mean (SD) | 3.4 (±0.5) | 3.6 (±0.5) | 0.11 |

Male FD patients (n=20, mean age 50.4 ±10.7 years) showed a more severe clinical phenotype than female FD patients (n=43, mean age 47.4 ±18.2 years). Frequency of LVH (37.2 % vs. 70.0 %, p = 0.029), CNVD (13.9 % vs. 55.0 %, p = 0.002), and heart valve disease (HVD, 0% vs. 15.0%, p = 0.028) was significantly higher in male patients. Consequently, the ADS3 was significantly higher (7.7 vs. 11.7, p = 0.005), and male FD patients showed significantly higher levels of LysoGb3 (4.2 vs. 27.0, p = 0.028). CV risk factors and age were not significantly different between male and female FD patients. Ferritin was significantly elevated in male FD patients (**Supplementary Table 3**).

### Impaired retinal microvascular function in Fabry disease

FD patients demonstrated significantly lower venular flicker-induced dilation (vFID) compared to age- and gender-matched HC (vFID: 3.5 % ± 1.6 % vs. 4.6 % ± 2.4 %; p = 0.006). No significant difference was observed in arteriolar flicker-induced dilation (aFID) between the groups, although FD patients showed a tendency towards higher values (3.9 % ± 3.1 % vs. 3.0 % ± 2.0 %; p = 0.08) (**Figure 1 A and B**). We analyzed retinal fundus pictures to assess retinal vessel diameters in FD patients. FD patients showed significantly narrower retinal arterioles (CRAE) when compared with HC (165.2 µm [153.5 - 183.2] vs. 183.2 µm [174.7 - 191.4], p < 0.001). There were no differences in retinal venular diameters (CRVE) between the groups (207.2 µm [196.9 - 217.2] vs. 212.6 µm [200.2 - 219.5]; p= 0.35). The arteriolar-venular ratio (AVR), calculated as CRAE/CRVE, was significantly lower in FD patients compared to HC (0.82 [0.74 - 0.87] vs. 0.86 [0.82 - 0.91]; p < 0.001) (**Figure 1 C, D and E**).

**Figure 1.**
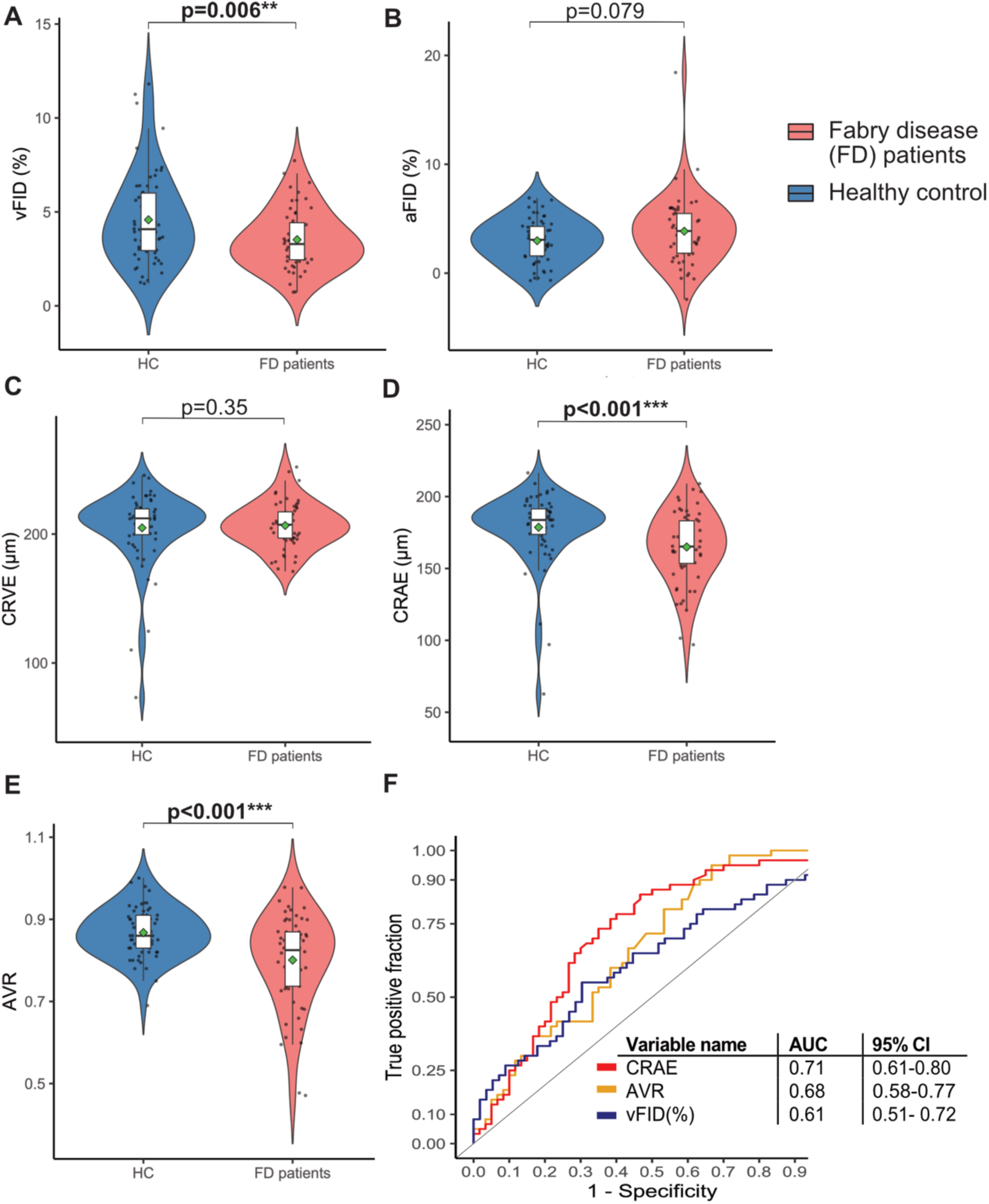
Dynamic retinal vessel parameters and static retinal vessel parameters in Fabry disease patients compared to healthy control. Violin plots of DVA parameters venular flicker-induced dilation (vFID: FD, n=56; HC, n=60; A) and arteriolar flicker-induced dilation (aFID: FD, n=55; HC, n=60; B) and SVA parameters CRVE (C), CRAE (D), and AVR (E; all n=60) in age- and gender-matched Fabry patients (red) and healthy controls (blue). Boxplots display values as the median (line) and mean (green square). The Wilcoxon rank-sum test was used for non-parametric distributions, and the Student’s t-test was used for parametric distributions. Statistical significance is indicated as follows: *p < 0.05; **p < 0.01; ***p < 0.001. ROC curves for CRAE, AVR and vFID and their AUCs and 95% confidence intervals (CI) are shown (F).

After controlling for potential confounders of RVA parameters, lower CRAE (p = 0.004), lower AVR (p < 0.001), and lower vFID (p = 0.002) remained significantly associated with FD (**Supplementary Table 4**). Additionally, vFID, CRAE, and AVR remained significantly lower in FD patients compared to HC after the exclusion of FD patients with a likely benign variant or VUS (**Supplementary Figure 1**). CRAE had good accuracy in distinguishing FD patients from HCs, with an area under the curve (AUC) value of 0.71. Both vFID and AVR achieved acceptable discrimination with AUC values of 0.61 and 0.68, respectively (**Figure 1 F**)

FD is an X-linked inherited disease; therefore, we conducted a subgroup analysis. For DVA and SVA parameters, we saw similar trends for both genders. Female FD patients had lower vFID (p=0.030) as did male patients, which, however, failed to reach significance (p=0.10). Male patients exhibited a significant increase in aFID compared to HCs (p = 0.027), whereas no significant difference was found in female FD patients (p = 0.72) (**Supplementary Figure 2 A and B**). CRVE did not differ significantly between female or male FD patients and HCs. CRAE and AVR were lower in male and female FD patients than HCs, with AVR not reaching significance in female patients (**Supplementary Figure 2 C, D, and E**). Thus, retinal microvascular changes are evident in male and female FD patients.

### Retinal vessel diameters and function as markers for clinical outcome in Fabry disease

Microvascular changes and their associated complications drive comorbidities and symptom severity in FD patients. Symptom severity in FD patients was assessed using the ADS3 score. Higher LysoGb3 levels were positively correlated with ASD3 (r = 0.26, p = 0.042). CRAE showed a strong negative correlation with symptom severity (r = - 0.40, p = 0.0013) (**Figure 2 A** and **B**). CRVE showed no significant correlation with ADS3 (r = −0.16, p = 0.22). However, lower AVR was significantly correlated with higher ADS3 (r = −0.33, p= 0.0098) (**Supplementary Figure 3 A and B**). DVA parameters showed no association with ADS3 (**not shown**).

**Figure 2.**
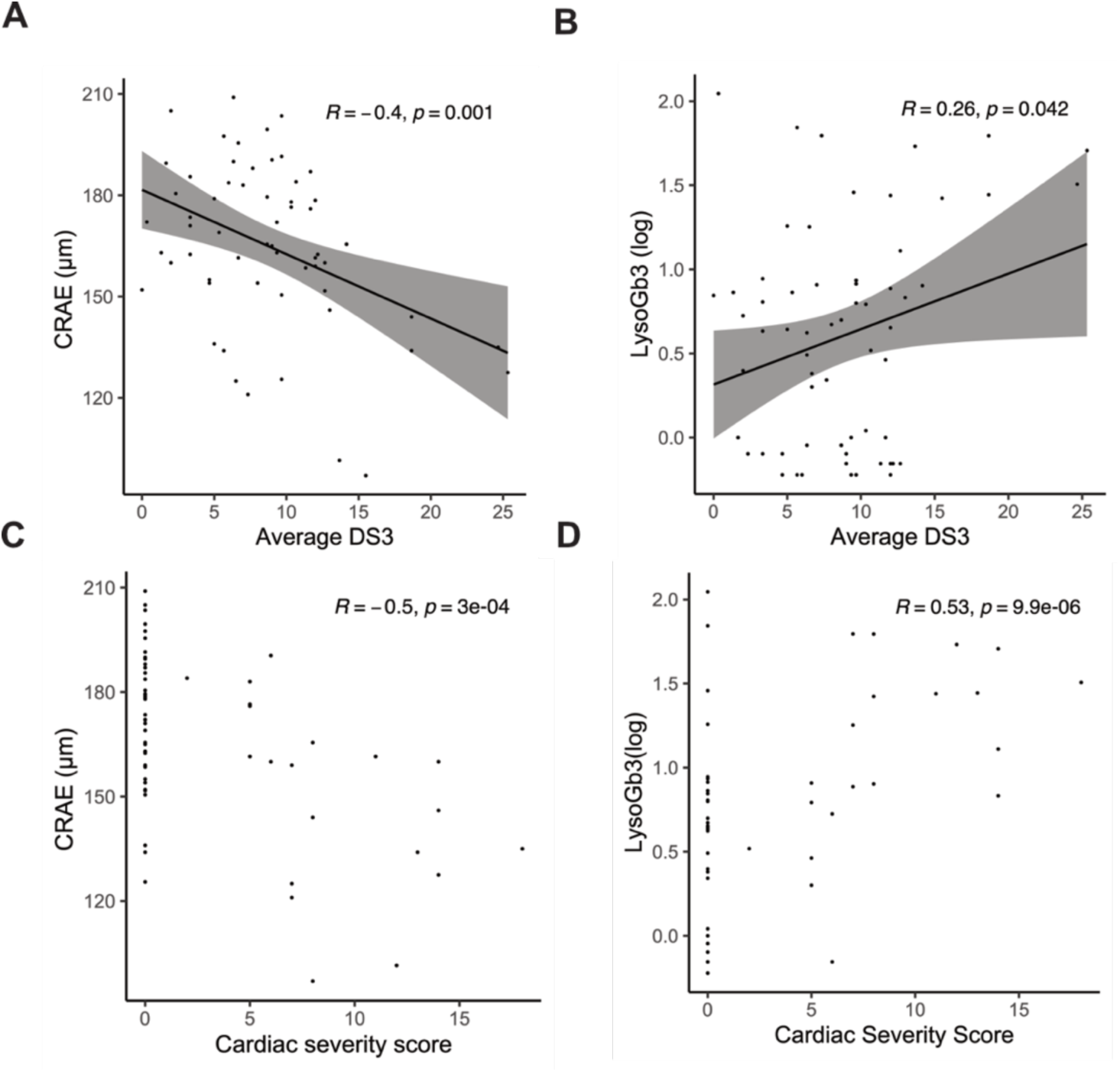
Correlation between altered retinal microcirculation and LysoGb3 with Average DS3 and Cardiac involvement. Scatterplots show the Pearson correlations between CRAE (n=60) or LysoGb3 (log-transformed, n=63) with Average DS3 (n=63), including the corresponding correlation coefficient (r) and p-value (A and B). Spearman correlations are shown for CRAE or LysoGb3 with the Cardiac Severity Score (C and D).

Among the four clinical domains of the ADS3, cardiac symptom severity was strongly correlated with both elevated LysoGb3 (r = 0.52, p<0.001) levels and narrower retinal arterioles (r = −0.5, p<0.001) (**Figure 2 C and D**).

Multivariable linear regression models were fitted to further explore the association between the retinal microcirculation and the cardiac phenotype. Lower CRAE and higher LysoGb3 levels were significantly associated with higher ADS3, which remained significant after adjusting for CV risk factors (**Table 2**). Narrow retinal arterioles were independently linked to specific organ involvement, showing significant associations with LVH, HVD, CNVD, and CKD. Higher LysoGb3 levels were mainly associated with LVH and HVD **(Table 2).** Similar findings were observed for lower AVR and CRVE, which also showed significant associations with increased cardiac symptom severity and organ involvement **(Supplementary Table 5).**

**Table 2.**
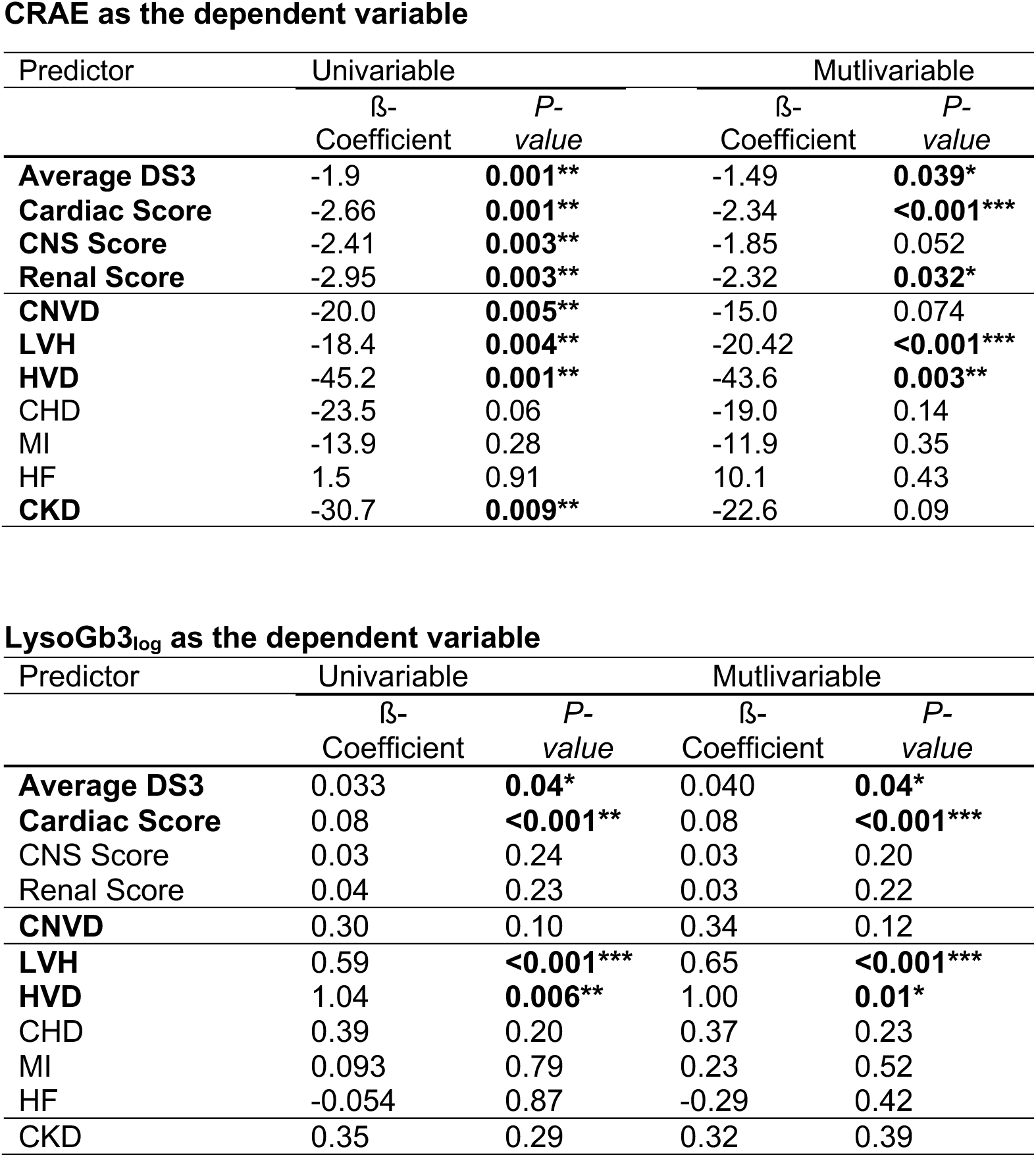
Associations between CRAE and LysoGb3, disease severity scores and cardiac symptom burden. Linear regressions with ß-Coefficient and the p-value are shown for CRAE (n=60) and LysoGb3 (n=63) for different predictor variables. A multivariable model was fitted for potential confounders: arterial hypertension, hypercholesterolemia, nicotine abuse, and BMI. CNS, central nervous system; CNVD, central nervous vascular disease; LVH; left ventricular hypertrophy; HVD, heart valve disease; CHD, coronary heart disease; MI, myocardial infarction; HF, heart failure; CKD, chronic kidney disease; LysoGb3 was log-transformed.

CRAE as the dependent variable
| Predictor | Univariable |  | Multivariable |  |
| --- | --- | --- | --- | --- |
| | β-Coefficient | P-value | β-Coefficient | P-value |
| **Average DS3** | -1.9 | **0.001\*\*** | -1.49 | **0.039\*** |
| **Cardiac Score** | -2.66 | **0.001\*\*** | -2.34 | **<0.001\*\*\*** |
| **CNS Score** | -2.41 | **0.003\*\*** | -1.85 | 0.052 |
| **Renal Score** | -2.95 | **0.003\*\*** | -2.32 | **0.032\*** |
| **CNVD** | -20.0 | **0.005\*\*** | -15.0 | 0.074 |
| **LVH** | -18.4 | **0.004\*\*** | -20.42 | **<0.001\*\*\*** |
| **HVD** | -45.2 | **0.001\*\*** | -43.6 | **0.003\*\*** |
| CHD | -23.5 | 0.06 | -19.0 | 0.14 |
| MI | -13.9 | 0.28 | -11.9 | 0.35 |
| HF | 1.5 | 0.91 | 10.1 | 0.43 |
| **CKD** | -30.7 | **0.009\*\*** | -22.6 | 0.09 |

**Table 2 Associations between CRAE and LysoGb3, disease severity scores and cardiac symptom burden**
| Predictor | Univariable |  | Multivariable |  |
| --- | --- | --- | --- | --- |
| | β-Coefficient | P-value | β-Coefficient | P-value |
| **Average DS3** | 0.033 | **0.04\*** | 0.040 | **0.04\*** |
| **Cardiac Score** | 0.08 | **<0.001\*\*** | 0.08 | **<0.001\*\*\*** |
| CNS Score | 0.03 | 0.24 | 0.03 | 0.20 |
| Renal Score | 0.04 | 0.23 | 0.03 | 0.22 |
| **CNVD** | 0.30 | 0.10 | 0.34 | 0.12 |
| **LVH** | 0.59 | **<0.001\*\*\*** | 0.65 | **<0.001\*\*\*** |
| **HVD** | 1.04 | **0.006\*\*** | 1.00 | **0.01\*** |
| CHD | 0.39 | 0.20 | 0.37 | 0.23 |
| MI | 0.093 | 0.79 | 0.23 | 0.52 |
| HF | -0.054 | 0.87 | -0.29 | 0.42 |
| CKD | 0.35 | 0.29 | 0.32 | 0.39 |

Given that static retinal vessel parameters and higher LysoGb3 levels were linked to cardiac symptom scores and comorbidities, we further investigated how SVA parameters correlate with cardiac laboratory markers and echocardiographic parameters. Lower CRAE was significantly associated with elevated levels of lactate dehydrogenase (LDH, r = −0.31), high sensitivity Troponin T (hs-Troponin T, r = −0.49) and NT-proBNP (r = −0.54). Lower AVR showed similar results. Regarding echocardiographic changes, both lower CRAE and AVR were significantly negatively correlated with the left ventricular end-diastolic diameter (LVEDD, r = −0.17 and r = - 0.19), intraventricular septum thickness (IVS, R=-0.55 and r = −0.38), and posterior wall thickness (PW, r = −0.46 and r = −0.32) (**Figure 3 A and C)**. Except for the correlation with LVEDD, similar correlations were observed for LysoGb3 (**Figure 3 B**). Lower CRVE was correlated with higher IVS (r = −0.3), PW (r = −0.28), and LDH (r = - 0.3) **(Figure 3 D).**

**Figure 3.**
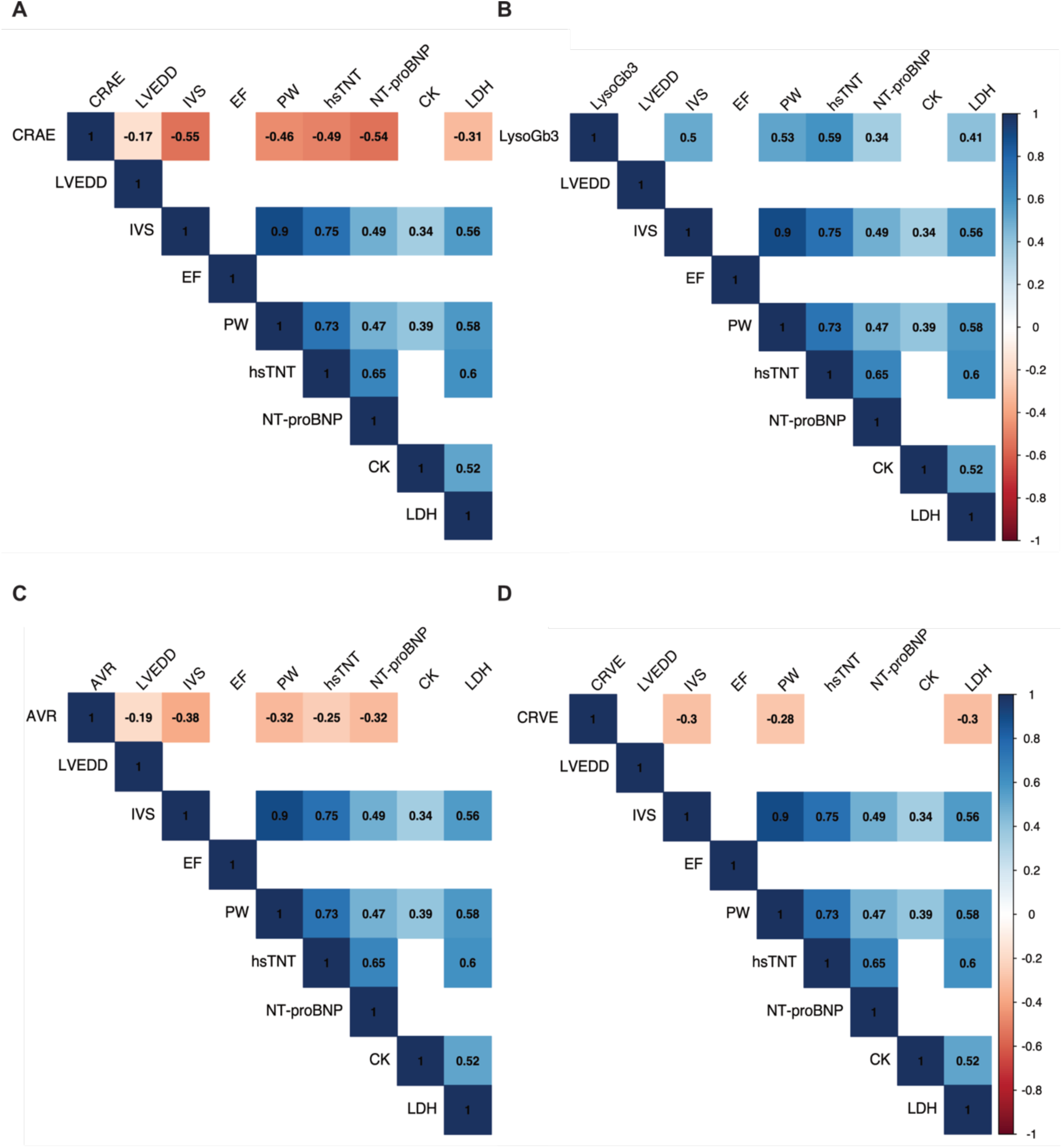
Correlations of static retinal vessel parameters and LysoGb3 with echocardiographic and laboratory parameters. Correlation plots show Spearman correlations of static retinal vessel parameters (A,C and D) and LysoGb3 (B) with cardiac laboratory parameters (hsTnT, high-sensitivity Troponin T, n=38; NT-proBNP, N-terminal prohormone of brain natriuretic peptide, n=38; CK, creatine kinase, n=63; LDH, lactate dehydrogenase, n=63) and echocardiographic parameters (LVEDD, left ventricular end-diastolic diameter, n=59; IVS, intraventricular septum thickness, n=60; PW, posterior wall thickness, n=58; EF, ejection fraction, n=46). Correlation coefficients (r) are visualized and displayed on the right. Non-significant correlations are not shown (empty fields).

### Retinal vessel health in Fabry disease patients treated with ERT

As we observed associations between an altered retinal microcirculation and disease severity, we were interested in whether ERT would influence retinal vessel health.

To address the bias that FD patients with higher Averaged DS3 and higher levels of LysoGb3 were more frequently treated with ERT in our cohort, we compared RVA parameters in FD patients stratified by symptom severity (high symptom score defined as ADS3 > median, and low symptom score as ADS3 < median) and treatment status (with or without enzyme replacement therapy). FD patients with lower disease severity showed stable CRAE values under ERT; in contrast, FD patients with high severity showed a tendency towards lower CRAE values under ERT, however, not significantly (p = 0.19) (**Supplementary Figure 4 A**). Other dynamic as well as static retinal vessel parameters showed similar results (**Supplementary Figure 4 B-E**). We fitted an interaction model to study the effect of disease severity on the association between CRAE and ERT. The interaction plot showed a significant interaction effect between ERT and ADS3 (p_interaction_ = 0.009), indicating that the effect of enzyme replacement therapy on CRAE depends on disease severity (**Supplementary Figure 4 F).**

### Fabry genetic variants, cardiac involvement and associations with retinal microvascular health

Sequence variants were classified according to the American College of Medical Genetics and Genomics (ACMG) guidelines, and clinical parameters were compared among four variant categories: pathogenic (n = 33), likely pathogenic (n = 7, variants of uncertain significance (VUS, n = 8), and likely benign (n = 14). There was a trend towards a higher frequency of Fabry-related comorbidities in patients with pathogenic, likely pathogenic, and VUS, though this did not reach statistical significance. High sensitivity (hs)-CRP and hs-TnT levels were significantly different across these clusters, with higher hs-CRP values observed in patients with VUS and pathogenic variants and higher hs-TnT levels in patients with likely pathogenic and pathogenic variants (**Supplementary Table 6**).

Because left ventricular hypertrophy is one of the most frequent signs of Fabry-related cardiomyopathy, we analyzed the relationship between IVS thickness, variant classification, and specific GLA variants.

In a multivariable linear regression model, IVS and LysoGb3 were significantly higher in VUS, likely pathogenic and pathogenic variants. Lower CRAE was associated with likely pathogenic and pathogenic variants (**Supplementary Table 7**).

Next, we assessed the relationship between specific GLA-gene sequence variants and IVS thickness in a multivariable model. Seven gene variants with a potential cardiac phenotype were selected based on their estimates and significant association with higher IVS, suggesting their relevance in FD-related cardiac hypertrophy (**Supplementary Table 8**).

We plotted a heatmap of scaled estimates for various cardiac outcome variables to further visualize potential cardiovascular involvement in the seven selected gene variants. Except for one variant (c.547G>A), all selected gene variants showed associations with either levels of higher cardiac laboratory parameters (LDH, CK, NT-proBNP, hsTnT) or higher LVEDD and PW thickness (**Figure 4**).

**Figure 4.**
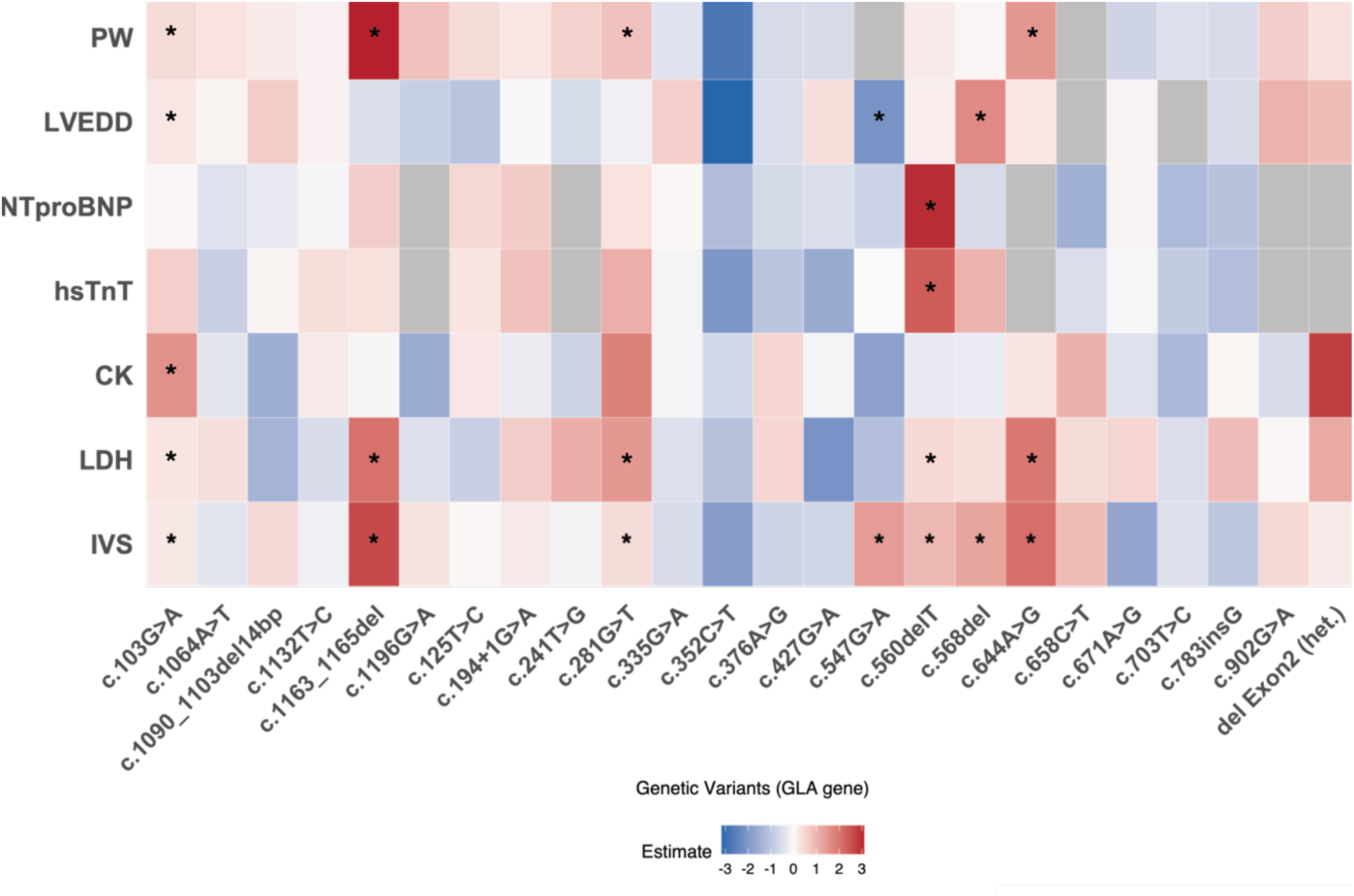
Heatmap of scaled estimates across GLA variants for cardiac outcome variables. This heatmap visualizes the scaled estimates of various cardiac outcome variables (y-axis) across GLA variants (x-axis, n=62). The color gradient represents the magnitude of the effect, with red indicating positive scaled estimates and blue indicating negative ones. Neutral estimates are shown in white; grey values indicate missing values. The outcome variables included IVS (n=60), LDH (n=62), CK (n=63), hsTnT (n=38), NT-proBNP (n=38), LVEDD (n=59) and PW (n=58). Statistically significant associations between GLA variants and IVS after correction for age, gender, and CV risk factors in a multivariable linear model are marked with asterisks in the first row. Additionally, significant associations between the potential seven cardiac variants and outcome variables after correction for age, gender, and CV risk factors in a multivariable linear model are marked with asterisks. The color bar shows the range of scaled estimates (ranging from negative=blue to positive=red), with deeper colors corresponding to stronger associations.

Next, we compared parameters of retinal microcirculation and LysoGb3 between FD patients with the six selected variants with a potential cardiac phenotype (n = 20) and FD patients with other GLA-gene sequence variants (n = 42). FD patients with a potential cardiac phenotype showed a significant increase in LysoGb3 levels (log-transformed, 0.81 vs. 0.02, p = 0.004). Additionally, they showed significantly narrower CRAE (159.5 µm vs. 171.0 µm, p = 0.039), indicating retinal microvascular dysfunction. Both CRVE and AVR showed a tendency towards lower levels, but not significant (**Figure 5 A-D**). Showcasing, cardiac involvement, NT-proBNP, and hsTnT were significantly higher in the selected six gene variants (**Figure 5 E and F**). Four of the variants with a potential cardiac phenotype were pathogenic, one likely pathogenic, and interestingly, one was a VUS. The selected gene variants were prominent in female FD patients (15/20 vs. 5/20).

**Figure 5.**
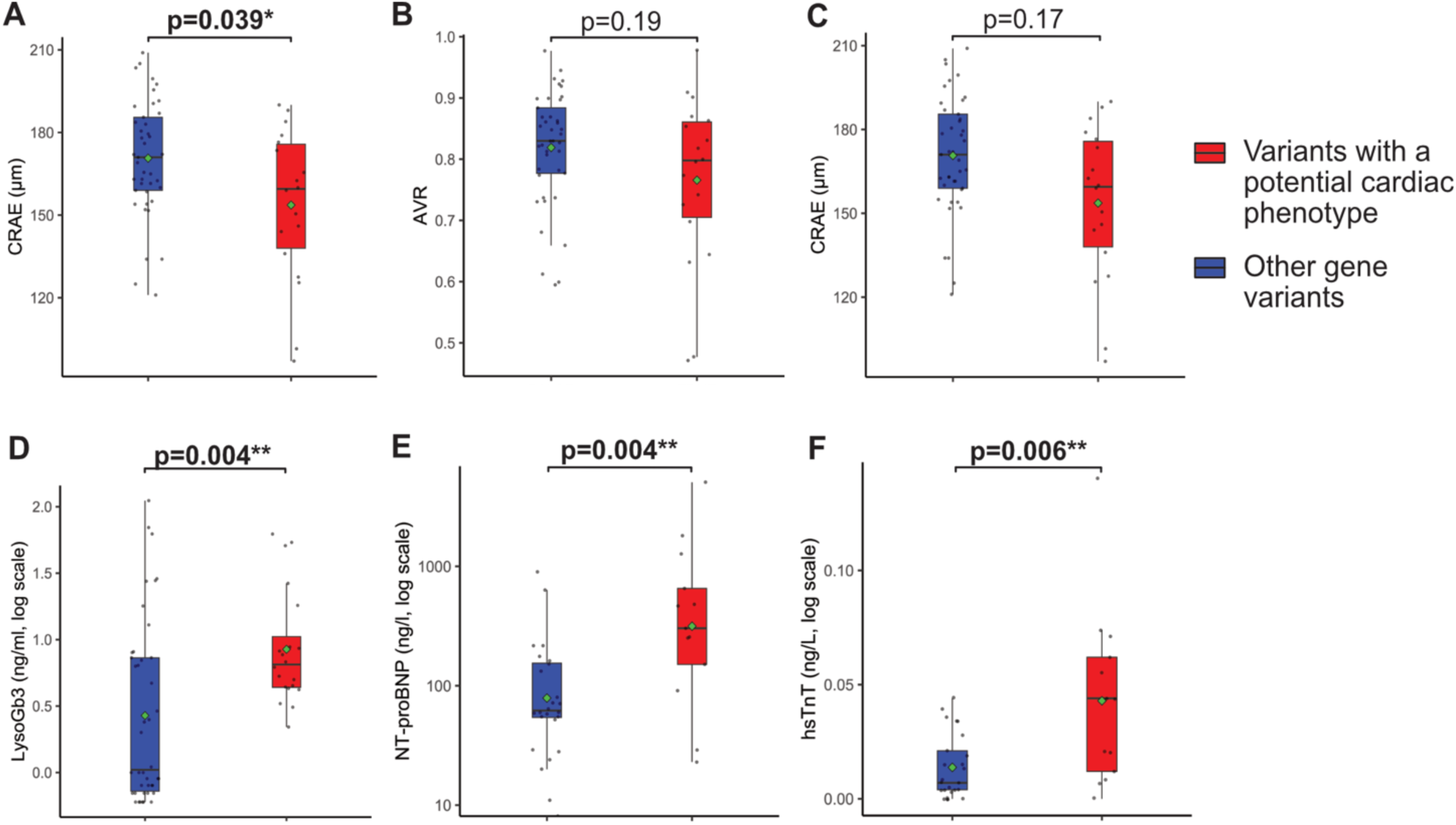
Retinal vessel parameters, LysoGb3 and cardiac laboratory values in FD patients with and without a potential cardiac variant. Boxplot of CRAE (A), AVR (B) and CRVE (C; all n=60), LysoGb3 (D; n=63), and laboratory parameters NT-proBNP (E; n=38) and hsTnT (F; n=38) between FD patients with a potential cardiac variant (red, n=20) and FD patients with other gene variants (blue, n=43). LysoGb3, hsTnT and NT-proBNP were log-transformed. Boxplot displays values as median (line) and mean (green square). The Wilcoxon rank-sum test was used for non-parametric distributions, and the Student’s t-test was used for parametric distributions. * p<0.05; **p<0.01; ***p<0.001.

### Inflammatory parameters and markers of endothelial dysfunction in FD patients

Previous studies have reported elevated markers of endothelial activation and chronic inflammation in FD patients. Compared to age- and gender-matched HC, FD patients displayed significantly elevated levels of VEGF (pg/ml, 67.3 [39.5 - 118.0] vs. 0.7 [0.0 - 2.1], p<0.001), ICAM-1 (ng/ml, 159.5 [78.6 – 333.6] vs. 0.0 [0.0 – 244.1], p = 0.039) and VCAM-1 (ng/ml, 48.9 [30.9 – 68.0] vs. 0 [0.0 – 25.2], p = 0.0011). In addition, levels of CXCL10 (pg/ml, 79.3 [45.9 - 120.3] vs. 37.5 [29.2 - 59.6], p<0.001), MCP1 (pg/ml, 94.9 [63.4– 117.7] vs. 37.0 [26.2– 65.0], p<0.001) and Rantes (ng/ml, 50.1 [15.0 – 125.6] vs. 1.4 [0.8 – 1.6], p<0.001) were elevated, IL-6 showed a tendency towards higher levels, however failing to reach significance (**Supplementary Figure 5 A - F**). All observed associations, except for lower ICAM-1 in FD patients, remained significant after controlling for potential confounders in a multivariable linear model (**Supplementary Table 9**).

We fitted regression models with interaction effects to investigate the association of markers of ED and inflammation markers with retinal vessel diameters and function in FD patients with a potential cardiac gene variant (**Supplementary Table 2**).

In FD patients with cardiac variants, the association between higher levels of CXCL10 and lower CRAE was stronger, with a significant interaction observed between CXCL10 and cardiac variants (p_interaction_=0.048) (**Figure 6 A**). Similar trends were found in FD patients with cardiac variants for the association between higher levels of MCP1, Rantes, and VCAM-1 and lower CRAE, although these interactions failed to reach significance (**Figure 6 B - D**).

**Figure 6.**
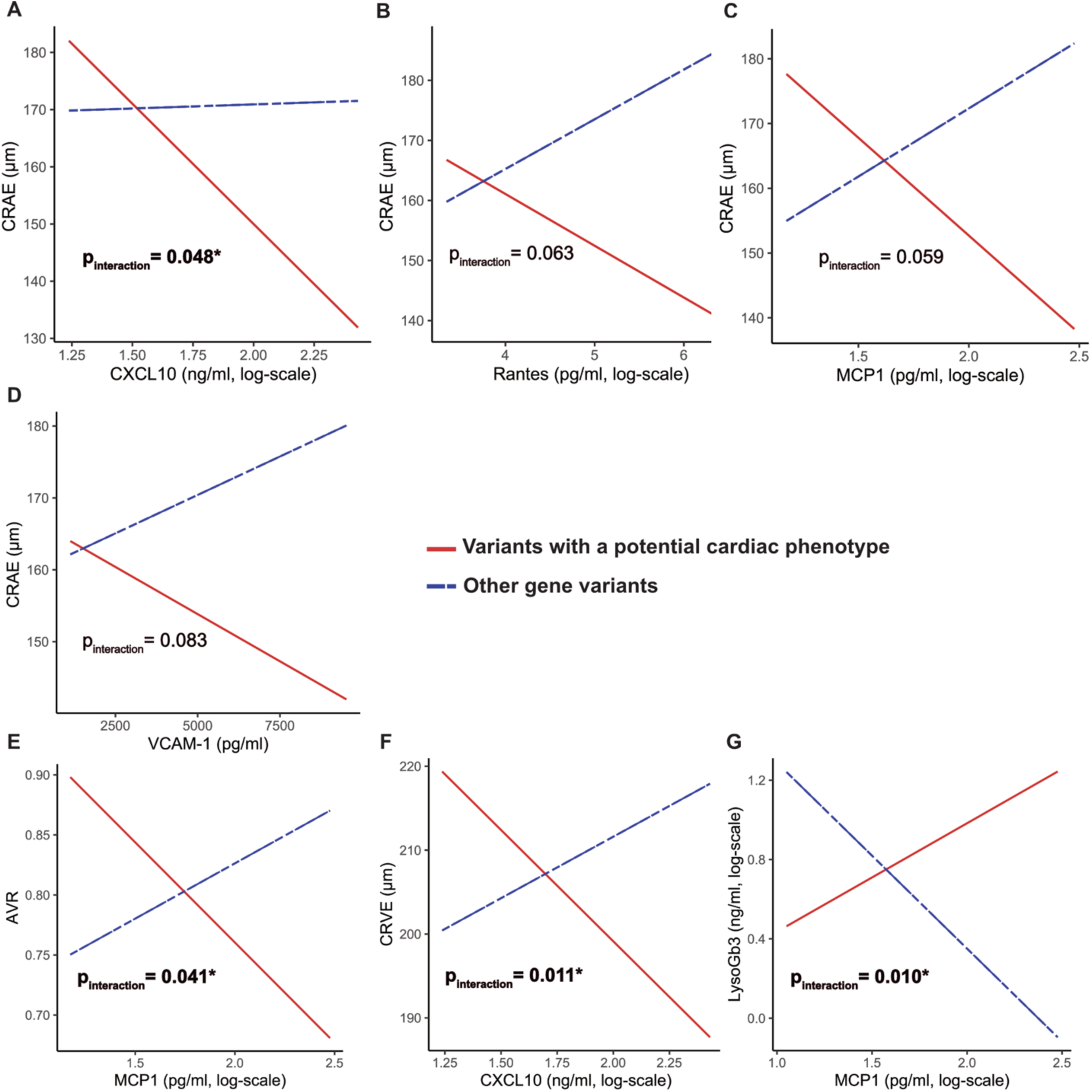
Interaction of endothelial dysfunction and inflammatory markers with static retinal vessel parameters and LysoGb3 in FD patients with cardiac gene variants. Interaction plots visualize the effect of cardiac gene variants (red line, n=20) and no cardiac variants (blue, dashed line, n=42) on the correlation between CRAE (A-D), AVR (E), CRVE (F), and LysoGB3 (G) with levels of inflammatory and endothelial dysfunction parameters. MCP1, CXCL10, Rantes and LysoGb3 were log-transformed (all n= 63). P_interaction_ displays the interactions between FD patients with a potential cardiac gene variant and laboratory variables, calculated in a multivariable linear model.

Consistent with these findings, in FD patients with cardiac variants, we observed a stronger association between higher CXCL10 levels and lower CRVE (p_interaction_= 0.011) as well as between higher MCP1 levels and lower AVR (p_interaction_= 0.041) (**Figure 6 E and F**). Interestingly, higher levels of LysoGb3 were associated with higher MCP1 in FD patients with cardiac variants (p_interaction_= 0.010) (**Figure 6 G**).

## Discussion

Endothelial dysfunction is a hallmark of Fabry disease, and cardiac impairment such as perfusion defects and slow coronary flow result from luminal narrowing in the microcirculation [10, 21, 38]. We could now show an altered retinal microcirculation in FD patients, as evidenced by significantly reduced flickering-induced venular dilatation, retinal arteriolar narrowing, and smaller AVR. Importantly, these microvascular differences were independent of age, gender, and CV risk factors such as arterial hypertension, BMI, hypercholesterolemia, and smoking.

Smooth muscle cell (SMC) proliferation is one of the consequences of GB3 deposition, and SMC proliferation is directly induced by LysoGb3 [19, 39]. As small arteries and arterioles in the retina are known to have relatively large components of SMCs, arteriolar narrowing, as evidenced by smaller CRAE in our FD patients, might be a direct effect of intima thickening [23]. SMC proliferation may lead to remodeling of the arterioles, increasing shear stress, which ultimately reduces NO synthesis. Studies also postulate a downregulation of the NO signaling pathway in FD [40, 41]. Both could explain our findings of reduced flicker-induced venular dilation. As we have shown that reduced vFID is an independent predictor of mortality in a cohort of patients with end-stage renal disease (ERDS), assessing venular dilation in FD patients could also provide valuable predictive insights, particularly in identifying patients at higher risk of Fabry-associated complications, such as cardiac or cerebrovascular events [27]. Systemic, ongoing inflammation may serve as the unifying pathophysiological mechanism underlying reduced venular dilation in both conditions [3, 42].

Interestingly, in male FD patients, flickering-induced arteriolar dilation was significantly higher than in healthy males. This might be explained by a compensatory overregulation in arterioles and, therefore, a stronger reaction to flickering light. Altarescu et al. also showed a stronger arterial vasodilation in FD patients after administration of acetylcholine [40]. This is in line with findings in Alzheimeŕs disease (AD), where the levels of storage products directly correlated with a higher arteriolar pulse amplitude, and the arteriolar pulse amplitude was significantly higher in AD patients [43, 44]. In female FD patients, CRAE and vFID were significantly reduced compared to HC, indicating that RVA provides a tool to quantify microcirculatory changes in female patients. In patients with residual enzyme activity, such as female patients or those with cardiac variants, SMCs showed deposits even when the endothelium remained clear. Therefore, SMCs may be more susceptible to Gb3 than the endothelium [45]. Monitoring female FD patients with RVA could provide additional insights into endothelial health beyond measuring often normal LysoGb3 levels. We argue that RVA could offer a cost-effective, non-invasive tool to monitor microvascular endothelial health in FD patients and, therefore, should be incorporated into routine care.

Whether ERT effectively clears Gb3 deposits in the heart remains controversial, and microvascular changes may persist well beyond therapy [16, 22, 46]. Our data suggest that ERT helps maintain or improve retinal arteriolar narrowing in FD patients with lower disease severity, potentially reflecting a therapeutic effect on the microcirculation. However, in patients with more advanced disease, ERT’s impact on CRAE is limited or altered, possibly due to longstanding disease-related vascular changes that are less responsive to ERT [6, 47]. One possible explanation could be persisting alterations in endothelial signaling pathways [17, 48]. Cell culture studies report an inherent defect in cholesterol trafficking, which may account for other long-term changes that ERT cannot reverse [49]. Additionally, secondary fibrosis, maintained by ongoing, unresolved inflammation, could impair ERT delivery to affected regions [16, 50].

Due to the increased availability of genetic testing and disease awareness, there is an evolving number of VUS [51]. Determining whether to start ERT can be challenging, especially with late-onset or organ-specific variants, such as cardiac ones [8, 52]. Although pathogenic variants tended to be associated with more Fabry-related complications in our cohort, complications were also observed in cases with VUS or even likely benign gene variants. After identifying six variants closely associated with primarily cardiac involvement during recruitment and predominantly found in female FD patients, we observed that these variants are strongly linked to arterial narrowing. This finding aligns with our observation that retinal arteriolar narrowing is closely associated with cardiac comorbidities, such as LVH, HVD, and CNVD, independent of traditional cardiovascular risk factors. Barbey et al. showed a proliferative effect of plasma from FD patients on SMC and cardiomyocytes, suggesting the plasma contains factors sustaining proliferative activity [53]. In our cohort, the patientś plasma showed elevated levels of chemokines, such as Rantes, MCP1, and CXCL10.

Rantes and MCP1 are upregulated in human podocytes after Lyso-Gb3 stimulation by the NOTCH-1-dependent pathway, contributing to podocyte injury and kidney fibrosis [54]. Additionally, in light of evidence linking the CXCL10/CXCR3 axis to proinflammatory and T-cell-mediated responses in cardiac diseases, our finding of elevated CXCL10 in patients suggests a potential role for CXCL10-mediated inflammation in the pathogenesis of cardiac complications in this cohort [55]. Chemokine-driven immune cell recruitment and amplified inflammatory signaling in endothelial cells may upregulate markers of endothelial activation in FD patients. We could show significantly higher levels of VEGF, ICAM-1, and VCAM-1. ICAM-1 and VCAM-1 are upregulated in EC after Gb3 stimulation, and VEGF has been associated with hypertrophic cardiomyopathy in FD patients [13, 56]. In FD patients with cardiac variants, we could now show that higher levels of MCP1 and CXLC10 are associated with an impaired retinal microcirculation and higher levels of LysoGb3. Cardiac inflammation and endothelial activation are secondary mechanisms sustaining damage to myocardial structures despite ERT [2, 3]. This further emphasizes how crucial early detection of microvascular changes in FD is and suggests that concomitant anti-inflammatory therapies should be considered in advanced disease. Higher levels of autoantibodies in FD patients further point toward a shared pathophysiology of autoinflammatory diseases and FD [57]. Further clinical studies are needed to target potential inflammatory pathways, such as the NLRP3 inflammasome and complement system, to stop an ongoing cycle of inflammation and secondary fibrosis [3, 50].

### Limitations of the Study

This study is cross-sectional; therefore, even though we tried to control for confounders, all correlations and associations should be interpreted as exploratory. Longitudinal studies are needed to confirm the predictive value of RVA parameters. While the study includes a relatively large cohort of 63 FD patients, which is significant given the rarity of the disease, subgroup analyses may still lack statistical power to detect certain associations. Furthermore, our cohort consisted primarily of heterozygous patients and we included FD patients with VUS and likely benign variants, and while subgroup analysis was performed, this composition may limit the generalizability of the findings.

Regarding the markers analyzed as secondary outcomes, it is essential to note that while the study was sufficiently powered to assess the primary outcome of endothelial dysfunction, the exploratory analyses involving laboratory markers and associations with genetic variants may not have been adequately powered to detect all possible associations. The results of these secondary analyses should be interpreted with caution and viewed as hypothesis-generating, providing a foundation for further studies with larger cohorts to validate these findings. Despite these limitations, this study represents a meaningful contribution to understanding endothelial dysfunction and its potential biomarkers in a rare disease like Fabry.

### Conclusion and outlook

Our study highlights the value of RVA as a quick, inexpensive, and validated tool for early detection of microvascular changes in Fabry disease, independent of traditional cardiovascular risk factors. The significant alterations observed in both static and dynamic retinal vessel parameters in male and female FD patients underline the close link between microvascular changes and systemic disease severity. Notably, the strong correlation between retinal arteriolar narrowing and established echocardiographic and laboratory markers of cardiovascular involvement suggests that RVA may be a reliable, non-invasive approach for monitoring endothelial health in Fabry disease patients.

Given the persistent inflammatory and endothelial activation seen in FD, particularly in patients with a potential cardiac gene variant, early detection and monitoring of microvascular abnormalities through RVA could enable more timely intervention and the possibility of incorporating adjunctive anti-inflammatory therapies to slow disease progression.

Longitudinal studies are essential to assess the predictive value of RVA for long-term cardiac- and Fabry-related outcome variables.

## Data Availability

Data are not available due to german data protections laws

## Acknowledgments

We want to thank all the patients who participated. Additionally, we want to thank our study nurse, Evi Altmann, and the doctoral students involved in the project: Tarek Assali, and Ahed Alachkar.

## Author Contribution Statement

CS, the Chief Investigator, and CR conceived the study and led the project. TK contributed to the study design, data interpretation and analysis, and drafting of the manuscript. ML and AR are lead methodologists regarding the basic science approach. RG helped develop the study and the manuscript. NH helped analyze the gene variants. HH, KK, and LS significantly helped with their expertise in RVA. All authors contributed to the manuscript development and accuracy of any part of the work. All authors read and approved the final manuscript.

## Supplementary material

Tables S1–S9

Figure S1–S5

## Non-standard Abbreviations and Acronyms

ADS3: Average Fabry Disease Severity Scoring System
aFID: Arteriolar Flicker-Induced Dilation
AVR: Arteriolar-Venular Ratio
CK: Creatine Kinase
CKD: Chronic Kidney Disease
CNVD: Central Nervous Vascular Disease
CRAE: Central Retinal Arteriolar Equivalent
CRVE: Central Retinal Venular Equivalent
CV: Cardiovascular
CVD: Cardiovascular Disease
CXCL10: C-X-C motif chemokine 10
DVA: Dynamic Retinal Vessel Analysis
EC: Endothelial Cell
ED: Endothelial Dysfunction
ERDS: End-Stage Renal Disease
ERT: Enzyme Replacement Therapy
FD: Fabry Disease
FMD: Flow-Mediated Dilation
Gb3: Globotriaosylceramide
HC: Healthy Control
HVD: Heart Valve Disease
ICAM-1: Intercellular Adhesion Molecule-1
IVS: Interventricular Septum
LDH: Lactate Dehydrogenase
LVEDD: Left Ventricular End-Diastolic Diameter
LVH: Left Ventricular Hypertrophy
LysoGb3: Globotriaosylsphingosin
MCP1: Monocyte Chemoattractant Protein-1
NLRP3: NOD-, LRR-, and Pyrin Domain-Containing Protein 3
NT-proBNP: N-terminal prohormone of Brain Natriuretic Peptide
PCT: Pharmacological Chaperone Therapy
PWV: Pulse-Wave Velocity
RANTES: Chemokine (C-C Motif) Ligand 5
RVA: Retinal Vessel Analysis
SMC: Smooth Muscle Cell
SVA: Static Retinal Vessel Analysis
VCAM-1: Vascular Cell Adhesion Molecule-1
VEGF: Vascular Endothelial Growth Factor
VUS: Variant of Uncertain Significance
vFID: Venular Flicker-Induced Dilation

